# Is There a Correlation Between Nitrate Quantity in Drinking Water and the Prevalence of Social Issues in the Metropolitan Area of Tehran?

**DOI:** 10.1101/2025.07.22.25332012

**Authors:** Habib Bagheri, Hossein Zamaninasab, Zohreh Bajelan, Milad Dehghan

## Abstract

**Background and Objectives:** Recent research by NGOs has revealed concerning nitrate levels in drinking water across all 22 districts of Tehran, amid conflicting reports from City Council officials and the Tehran Water and Waste Water Company regarding pollution. A preliminary study on groundwater in the U.S. demonstrated a strong link between nitrate levels and various societal issues. This study aims to investigate two key areas: the current nitrate levels in Tehran’s drinking water and their compliance with safety standards set by organizations like the WHO and US EPA. It also seeks to determine if there is a correlation between nitrate levels in specific Tehran districts and societal problems similar to those observed in the U.S.

**Materials and Methods:** Sampling took place on November 6, 2022, using 50 mL clear polypropylene Falcon test tubes, which were prepared by rinsing with distilled water. Twenty-two residential sites in Tehran’s districts were randomly chosen. Samples were collected within a 16-hour window, allowing the water to flow for 30 seconds before collection. The samples were immediately frozen at −4 degrees Celsius and analyzed 24 hours later for nitrate nitrogen using the colorimetric method (Brucine) as per U.S. EPA guidelines. The analysis was conducted at the Aryan Fan Azma laboratory using a portable DR 2800 spectrophotometer. Environmental conditions during testing included a temperature of 23.4 degrees Celsius and humidity of 18%. Statistical analyses were performed using the Shapiro-Wilk test, Kruskal-Wallis test, and hierarchical cluster analysis (HCA) with R and ApexChart.JS software, while heat maps were created using ArcGIS Pro.

**Findings:** The nitrate concentration measurements across Tehran’s districts revealed levels exceeding the Maximum Contaminant Level (MCL) set by WHO in districts 9, 15, and 16, contradicting previous studies. Furthermore, a significant correlation was found between nitrate concentrations and various social issues, including rates of illiteracy, incidents of harassment by vagrants and drug addicts, feelings of insecurity at night, and the prevalence of vagrants and drug users.

**Conclusion:** This research highlights a notable link between nitrate levels in drinking water and social challenges in Tehran. The correlation between high nitrate concentrations and various societal issues suggests a need for further investigation into this relationship. Additionally, these findings underscore the importance of integrating environmental and social health considerations into urban planning and public policy initiatives.

## 1. INTRODUCTION

### 1.1. Groundwater as a Major Source of Drinking Water

In the last century, with the significant growth of the human population, the global demand for access to fresh and clean water has increased exponentially (Luqman and Al-Ansari, 2021). One of the most important sources of freshwater in the world is groundwater. It is estimated that the drinking water of two billion people worldwide is directly supplied from groundwater sources, and most of the water used in agriculture and the food preparation industry depends on groundwater (Centers for Disease Control and Prevention, 2024).

Another source of water originates from wetlands, lakes, and dams, which are classified as surface water, while the infiltration of rain and snow into the empty spaces between soil grains and rocks leads to the formation of groundwater.

### 1.2. Water Supply in Iran and the Capital City of Tehran

With a decline of 74 cubic kilometers between 2002 and 2015, based on basin-scale estimates (Ashraf et al., 2021), Iran’s groundwater is being depleted at an alarming rate. This 14-year depletion surpasses 92% of the 57-year depletion observed in the U.S. High Plains from 1950 to 2007.

According to the Islamic Republic News Agency in January 2020 (Bahrami, 2020), approximately 57% of Iran’s total water needs and 65% of its drinking water needs rely heavily on groundwater sources. This water is extracted through three main methods: springs (n = 173,452), wells (n = 518,959), and aqueducts (n = 41,011).

As the second most populous metropolis in the Middle East, with a metropolitan population exceeding 16 million, the situation in Tehran is even more concerning than in other cities (Brinkhoff, 2024). According to statistics from the Tehran Province Water and Wastewater Company, 85 million cubic meters of water are collected monthly in the city, of which about 71.7% comes from surface water (via the Karaj, Lar, Latian, Taleghan, and Mamloo dams) and 28.3% from groundwater (Hamshahrionline, 2022; Tehran Water and Wastewater Company, 2020). In dry years, groundwater can account for 60–70% of Tehran’s drinking water supply (Mohammadi et al., 2008; Sarmadi et al., 2021).

In this context, while the average per capita availability of freshwater is only one-third of the global standard (Omrani, 2013), a resident of Tehran consumes an average of 326 liters of water per day—126% more than the average daily water consumption of European Union citizens (Bloomberg, 2014; Earthscan, 2011).

### 1.3. Possible Hazardous Effects of Nitrates

One of the most significant potential pollutants in water resources is the nitrate ion. Under natural conditions as part of the nitrogen cycle, nitrate should constitute only a very small mass percentage of drinking water (Lin et al., 2023). However, issues such as the excessive use of fertilizers in agriculture (Komor and Anderson, 1993; Wassenaar, 1995) have led to a substantial increase in nitrate levels in both surface and groundwater in Iran (Jalali, 2005).

For decades, it was assumed that groundwater was safe from various pollutants. However, subsequent studies have shown that human activities significantly contribute to groundwater contamination, including petroleum products (Meyers, 2011), pesticides, sewage effluents (Wakida and Lerner, 2004; Lee et al., 2020), and many other chemicals.

According to the World Health Organization (WHO) and the U.S. Environmental Protection Agency (EPA), the maximum contaminant levels (MCLs) for nitrate ions in drinking water are 50.0 mg/L and 44.3 mg/L as nitrate, respectively—equivalent to 11.3 mg/L and 10 mg/L as nitrogen (Ibe et al., 2021; World Health Organization, 2003). These limits are primarily designed to prevent the most well-known health consequence of high nitrate exposure: neonatal methemoglobinemia (Li and Weng, 2007; Ward et al., 2018). However, other nitrate-related health risks have been overlooked in determining these MCLs. These include cancer, birth defects (Blaisdell et al., 2019), thyroid dysfunction (Eskiocak et al., 2005), and various pregnancy-related disorders such as spontaneous abortion and low birth weight (Ward et al., 2018).

More recently, academic studies have investigated the neurological effects of nitrate consumption in living organisms. For example, research has shown increased nitrite/nitrate concentrations in the prefrontal cortex of stressed subjects, in the hippocampus of 6-OHDA-treated animals, and in the striatum of those subjected to both stress and 6-OHDA. This increase in nitrite and nitrate is closely linked to nitrosative stress and elevated nitric oxide levels. Excessive nitric oxide production can react with superoxide anions to form peroxynitrite, a highly reactive oxidant. The overproduction of reactive nitrogen species (RNS) triggers inflammatory and oxidative processes, eventually leading to neurotoxicity. These effects are associated with dysfunction in the glutamatergic system, which plays a role in Parkinson’s disease (PD) and major depressive disorder (MDD). Elevated levels of nitric oxide and nitrate have been observed in individuals with MDD, while oxidative stress is also implicated in PD (Tuon et al., 2021).

Dietary nitrates also play a physiological role, as they are metabolized into nitric oxide (NO) through complex pathways. NO contributes to immune modulation, neurotransmission, and vasodilation. The vasodilatory effect enhances blood flow and oxygen delivery to tissues, thereby supporting cardiovascular health, exercise performance, and cognitive function. However, individual responses to dietary nitrates can vary (Apte et al., 2023). In studies on zebrafish, nitrate treatment reduced startle reactivity but preserved habituation. The fish showed delayed decision-making and longer reaction times in shuttle-box tasks, suggesting deficits in associative learning and executive function. Nitrate exposure may enhance cerebral blood flow and cognitive performance via NO-mediated vasodilation, but prior studies have produced inconsistent findings on nitrate’s effects on mood and cognition. Nitrate and nitrite treatments also reduced brain levels of GABA and glutamine, possibly contributing to anxiety- and depression-like behaviors, as the GABAergic system is closely tied to emotional regulation. Additionally, they influenced purine metabolism in ways that resemble mitochondrial dysfunction and aging. Nitrite exposure promoted fatty acid oxidation, potentially affecting brain development and function. Changes in membrane polyunsaturated fatty acids (PUFAs) could impact dopamine signaling and heighten the risk of neurological disorders (García-Jaramillo et al., 2020).

### 1.4. Nitrate Pollution in Tehran’s Drinking Water

Until 2021, Tehran’s untreated and hazardous urban wastewater was directly discharged into underground water sources (Tajrishy et al., 2014). With the development of an 8,000 km underground sewage network, this issue has been largely mitigated (Tehran Water and Wastewater Company, 2015). However, serious concerns remain regarding the safety of Tehran’s drinking water (Shargh, 2022). City officials have made contradictory statements about water quality. For example, two official organizations have reported that nitrate concentrations in some parts of Tehran are threatening and carcinogenic (Shafieian, 2014) or at worrying levels (Donya-e-Eqtesad, 2016), while other officials have claimed that the city’s drinking water is completely safe (Shafieian, 2014). Amid these contradictions, in recent years, several wells in southern Tehran have been shut down due to excessive nitrate concentrations (Shafieian, 2014; Borna, 2019).

A study conducted in 2009 by a university research team (Mohammadi et al., 2012) involved random sampling at 105 water points across Tehran. The findings indicated that the average nitrate concentration in the sampled areas was 18.09 mg/L, with a range from 2.65 to 87.5 mg/L.

### 1.5. Research Objectives

Like other densely populated cities around the world, Tehran is increasingly confronted with various social challenges (Jelokhani-Niaraki et al., 2019). The Nitrate-Crime Hypothesis (Zamaninasab and Bajelan, 2023) has proposed a potential link between the concentration of nitrate ions in drinking water and the prevalence of a broad range of social harms. In the present study, we measured nitrate ion concentrations in the drinking water of all 22 districts of Tehran. We then compared the results with both prior reports and the WHO’s maximum contaminant level (MCL) standards. Finally, we examined the relationship between the prevalence of various social problems and the nitrate levels in drinking water across the different districts of Tehran.

## 2. MATERIALS AND METHODS

### 2.1. Study Area

Tehran is situated on the southern slopes of the Alborz Mountains in northern Iran, between latitudes 35°30′ to 35°51′ N and longitudes 51°00′ to 51°40′ E (Sohrabinia and Khorshiddoust, 2007) (Figure 1).

**Figure 1.**
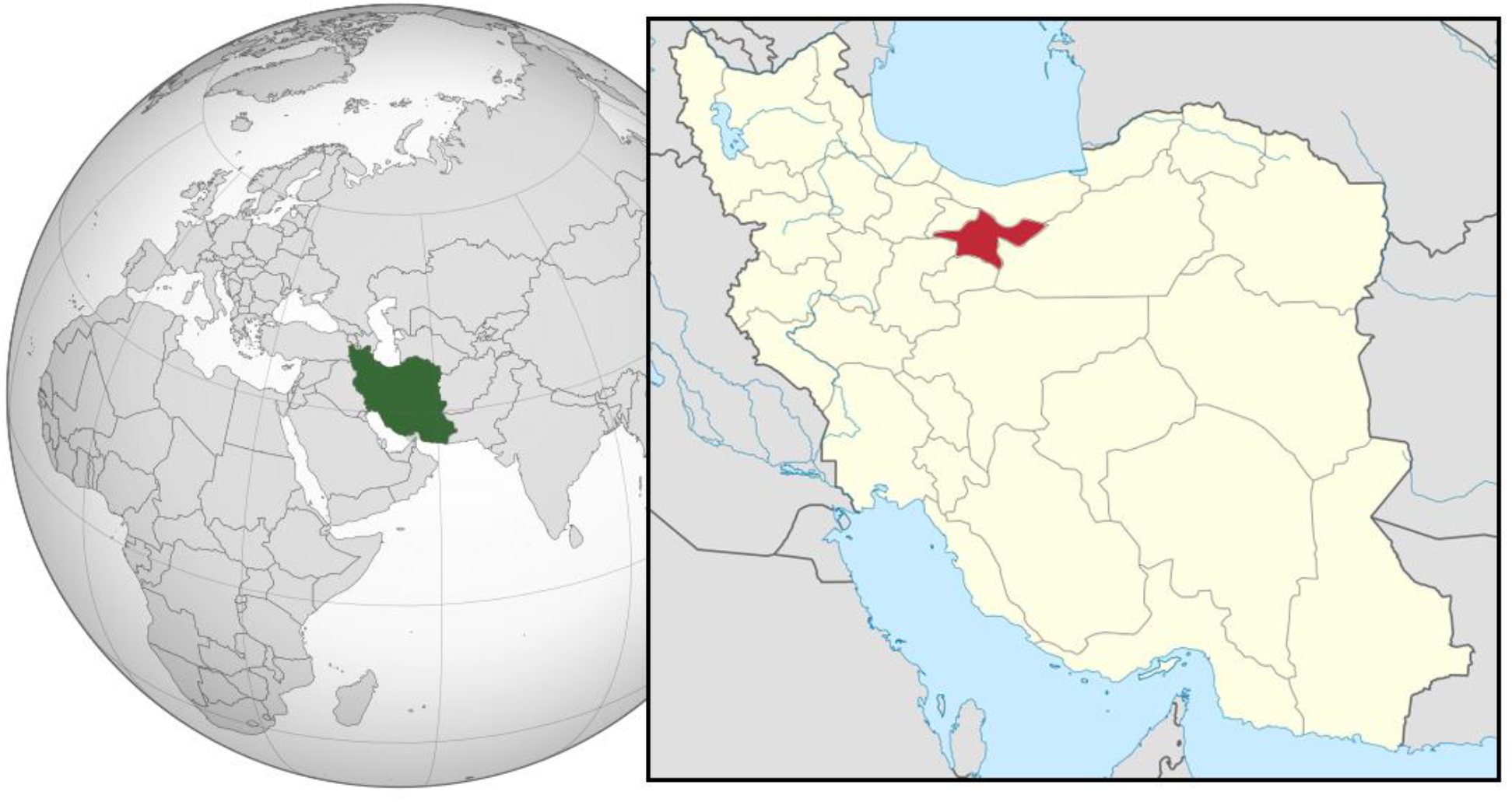
Tehran within Iran, and Iran within the world context (P30Carl, 2024).

The city is administratively divided into 22 municipal districts. Additionally, Tehran is segmented into six water and wastewater service zones—known as Tehran ABFA districts—within which these municipal districts are located (Figure 2).

**Figure 2.**
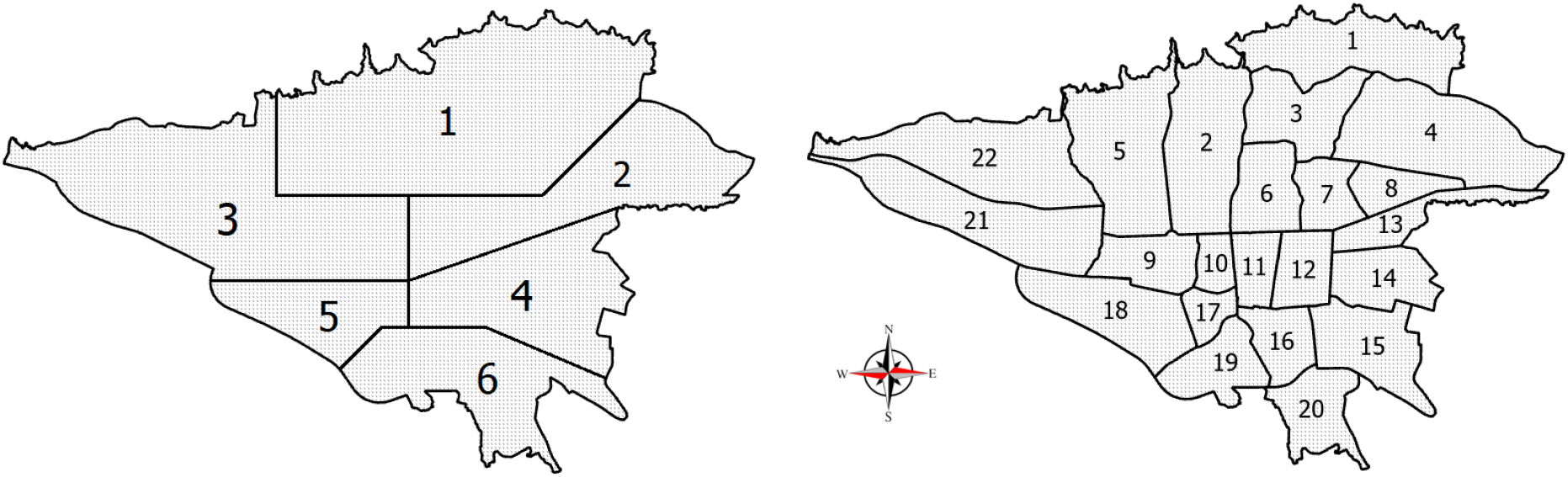
Approximate map of Tehran’s 22 municipal districts (right) and its six ABFA (water and wastewater service) districts (Tehran Water and Wastewater Company, 2020).

### 2.2. Collection of Samples

Sampling across all selected areas was conducted on a single day: Sunday, November 6, 2022. Water samples were collected in 50 mL conical test tubes made of clear polypropylene, which were both autoclavable and suitable for freezing. Prior to sampling, the tubes were rinsed with distilled water and air-dried.

Twenty-two residential locations—one from each of Tehran’s 22 municipal districts—were randomly selected (labeled A to V on the map in Figure 3). Over a 16-hour period (from 8:00 a.m. to 12:00 a.m. local time), 50 mL samples were taken from these points. In each case, the same standardized procedure was followed: the tap water was allowed to flow for 30 seconds before the sample was collected.

**Figure 3.**
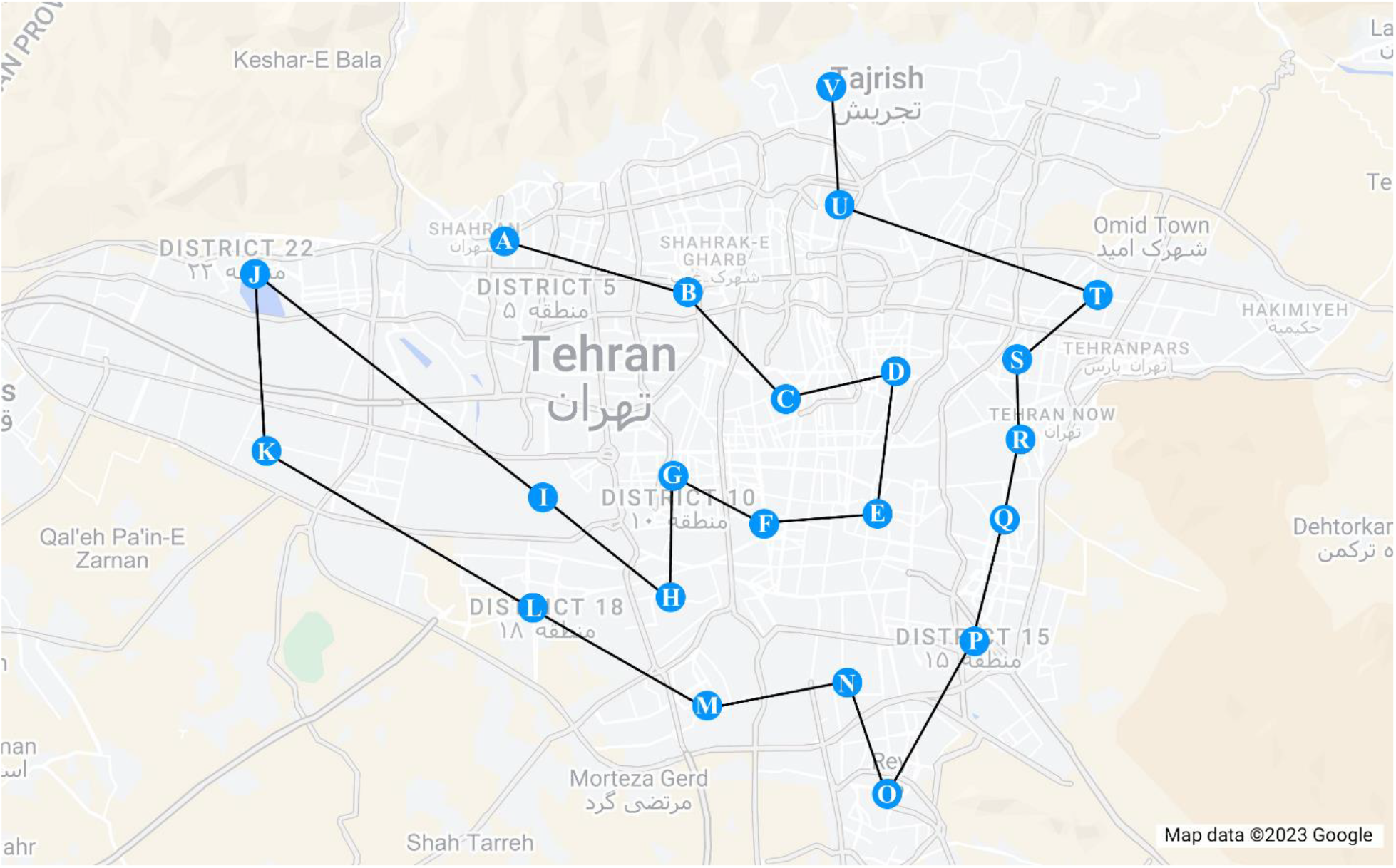
The route taken for sampling

Immediately after collection, all samples were stored in a portable freezer at –4°C to preserve their chemical integrity.

### 2.3. Nitrate Concentration Analysis

Twenty-four hours after sampling, the water samples were analyzed using Method 352.1: Nitrogen, Nitrate (Colorimetric, Brucine), as described by the U.S. Environmental Protection Agency (EPA, 1971). The analysis was performed via spectrophotometry at the Arian Fan Azma Laboratory (registration code: 0112277A) using the DR 2800™ Portable Spectrophotometer. Laboratory conditions during testing included a temperature of 23.4°C and a relative humidity of 18%.

Data tables were initially prepared in Microsoft® Excel® LTSC MSO 32-bit and subsequently imported into R software (version 4.2.2) for statistical analysis, graph generation, and Hierarchical Cluster Analysis (HCA). ArcGIS Pro (version 3.0.2) was employed to generate the spatial heat map of nitrate concentration.

The Shapiro–Wilk test was applied to assess the normality of nitrate distribution across Tehran’s 22 districts. Originally introduced in 1965 by Shapiro and Wilk (Shapiro and Wilk, 1965), this test evaluates the null hypothesis that a given sample is normally distributed. A small p-value indicates a rejection of the null hypothesis.

The Kruskal–Wallis test was used to examine whether the mean nitrate concentrations across the 22 municipal districts significantly differ when grouped by Tehran’s six water and wastewater (ABFA) service zones. This non-parametric test, proposed by Kruskal and Wallis in 1952, tests the null hypothesis that the group mean ranks are equal. A small p-value suggests a statistically significant difference (Xia, 2020).

Hierarchical Cluster Analysis (HCA) was used to classify the 22 districts based on the similarity of nitrate concentrations. This multivariate statistical method groups samples by minimizing intra-group variance and maximizing inter-group variance (Da Silva Torres et al., 2005).

### 2.4. Social Problems Studied

The primary sources of data on social issues in this study were the Atlas of Urban Quality of Life in Tehran Metropolis (Tehran Municipality, 2017)—a document prepared in collaboration with the Vice-Chancellor for Social and Cultural Affairs of Tehran Municipality and the University of Tehran—and a detailed report on social issues in the 22 districts published by the Social and Cultural Studies Office of the Municipality of Tehran (Deputy of Social and Cultural Affairs of Tehran Municipality, 2024) (Table 1).

**Table 1.** Social indices of the present study.

| # | Index | Type | Description |
| --- | --- | --- | --- |
| 1 | Being mugged | Ranking | This shows the ranking of regions based on the number of people who have been mugged (Tehran Municipality, 2017). |
| 2 | Vehicle theft | Ranking | This shows the ranking of districts based on vehicle theft (Tehran Municipality, 2017). |
| 3 | Social harms | Ranking | This shows the ranking of districts based on the social harms (Irankhah and Momeni, 2020) |
| 4 | Daily woman safety | Ranking | This shows the ranking of the districts based on the daily safety of women in the neighborhood (Tehran Municipality, 2017). |
| 5 | Security Feeling Index | Ranking | This shows the ranking of districts based on the neighborhood Security Feeling Index (Tehran Municipality, 2017). |
| 6 | Divorced or widowed | Prevalence | This shows the percentage of single people due to divorce or the death of a spouse in each district (Deputy of Social and Cultural Affairs of Tehran Municipality, 2024). |
| 7 | Illiteracy or low-literacy | Prevalence | This shows the percentage of people who are illiterate or have only primary education in each district (Deputy of Social and Cultural Affairs of Tehran Municipality, 2024) |
| 8 | Unemployment | Prevalence | This shows the percentage of unemployed people in each district (Deputy of Social and Cultural Affairs of Tehran Municipality, 2024). |
| 9 | Harassed by vagrants | Prevalence | This shows the percentage of people who were disturbed by neighborhood vagrants, a lot or somewhat, during the last year (by district) (Deputy of Social and Cultural Affairs of Tehran Municipality, 2024). |
| 10 | Harassed by addicts | Prevalence | This shows the percentage of people who were disturbed by neighborhood addicts, a lot or somewhat, during the last year (by district) (Deputy of Social and Cultural Affairs of Tehran Municipality, 2024). |
| 11 | Being pickpocketed | Prevalence | This shows the percentage of people who were pickpocketed in each district (Deputy of Social and Cultural Affairs of Tehran Municipality, 2024). |
| 12 | Being physically assaulted | Prevalence | This shows the percentage of people who were physically assaulted in their neighborhood in each district (Deputy of Social and Cultural Affairs of Tehran Municipality, 2024). |
| 13 | Being harassed | Prevalence | This shows the percentage of people who were harassed in their neighborhood in each district (Deputy of Social and Cultural Affairs of Tehran Municipality, 2024). |
| 14 | Not feeling safe at night | Prevalence | This shows the percentage of people who found it very/somewhat unsafe to walk in their neighborhood at night (by district) (Deputy of Social and Cultural Affairs of Tehran Municipality, 2024). |
| 15 | Addicts in neighborhood | Prevalence | This shows the percentage of people in each district who admitted to seeing drug addicts a lot/somewhat while passing by in their neighborhood (Deputy of Social and Cultural Affairs of Tehran Municipality, 2024). |
| 16 | Not feeling safe at home | Prevalence | This shows the percentage of people who described it as very/somewhat unsafe to be alone at home at night (by district) (Deputy of Social and Cultural Affairs of Tehran Municipality, 2024). |
| 17 | Number of vagrants | Prevalence | This shows the percentage of people in each district who admitted to seeing drug vagrants a lot/somewhat while passing by in their neighborhood (Deputy of Social and Cultural Affairs of Tehran Municipality, 2024). |

## 3. RESULTS AND DISSCUSION

### 3.1. Overview

The results of nitrate concentration measurements at randomly selected sampling points across the 22 districts of Tehran are presented in Table 2 and Figure 4. The data indicate that the nitrate concentration in the drinking water of Districts 15, 9, and 16 exceeds the World Health Organization (WHO) maximum contaminant level (MCL).

**Table 2.** Nitrate concentration in drinking water samples from 22 districts of Tehran (results are sorted in descending order).

| Sample No | Sampling Place (High nitrate districts) | Nitrate Concentration (mg.L<sup>-1</sup>) | Sample No | Sampling Place (Low nitrate districts) | Nitrate Concentration (mg.L<sup>-1</sup>) |
| --- | --- | --- | --- | --- | --- |
| 1 | Dist. 15 | 65.0 | 12 | Dist. 22 | 24.1 |
| 2 | Dist. 9 | 65.0 | 13 | Dist. 3 | 18.7 |
| 3 | Dist. 16 | 61.0 | 14 | Dist. 8 | 15.0 |
| 4 | Dist. 19 | 49.5 | 15 | Dist. 5 | 8.0 |
| 5 | Dist. 17 | 48.0 | 16 | Dist. 7 | 6.5 |
| 6 | Dist. 18 | 45.0 | 17 | Dist. 13 | 6.2 |
| 7 | Dist. 11 | 35.0 | 18 | Dist. 2 | 5.2 |
| 8 | Dist. 10 | 34.8 | 19 | Dist. 6 | 4.7 |
| 9 | Dist. 12 | 32.0 | 20 | Dist. 4 | 3.8 |
| 10 | Dist. 20 | 30.0 | 21 | Dist. 21 | 3.1 |
| 11 | Dist. 14 | 28.8 | 22 | Dist. 1 | 2.1 |

**Figure 4.**
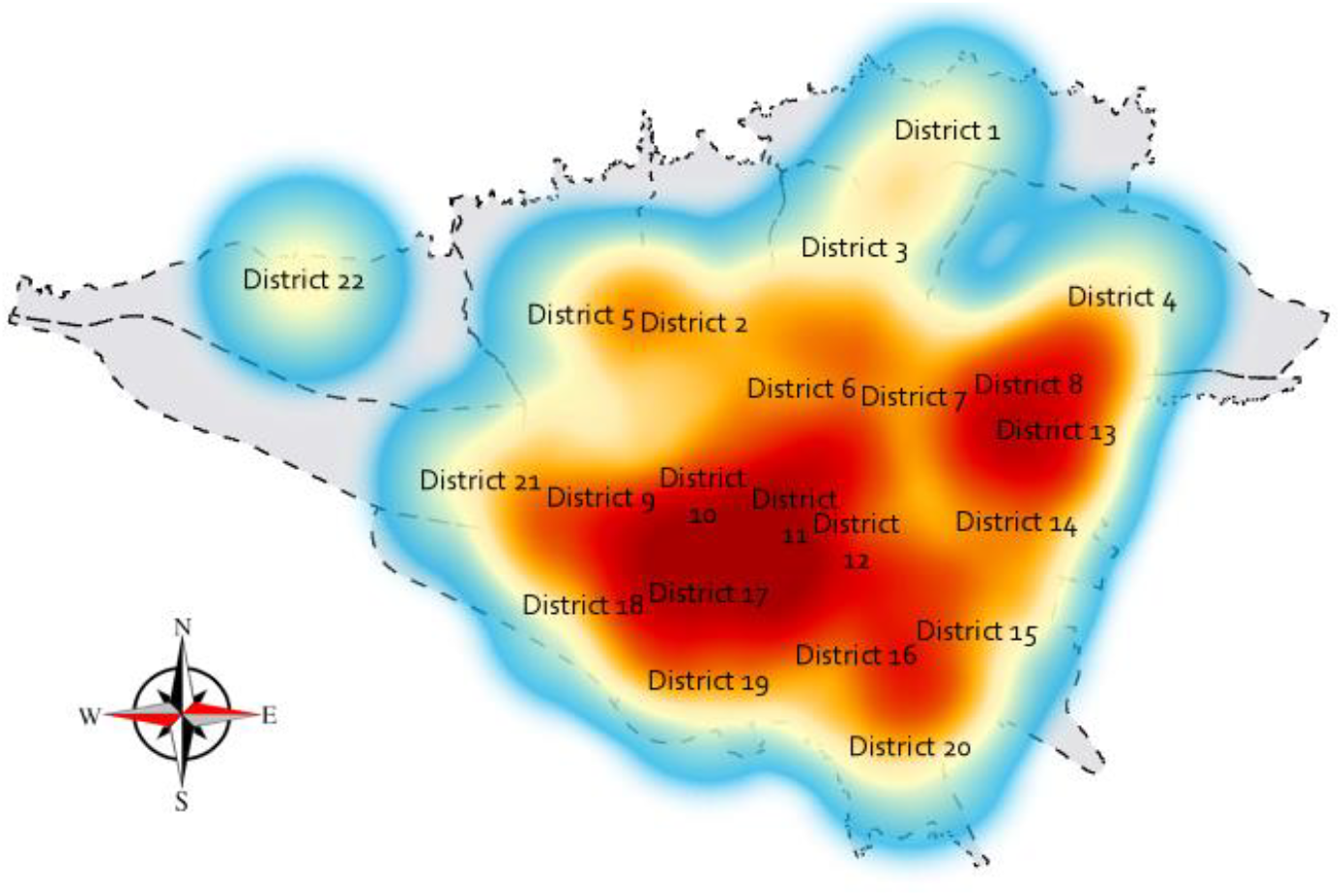
Heat map of nitrate distribution in drinking water in 22 districts of Tehran

The details of the statistical data are explained in Table 3.

**Table 3.**
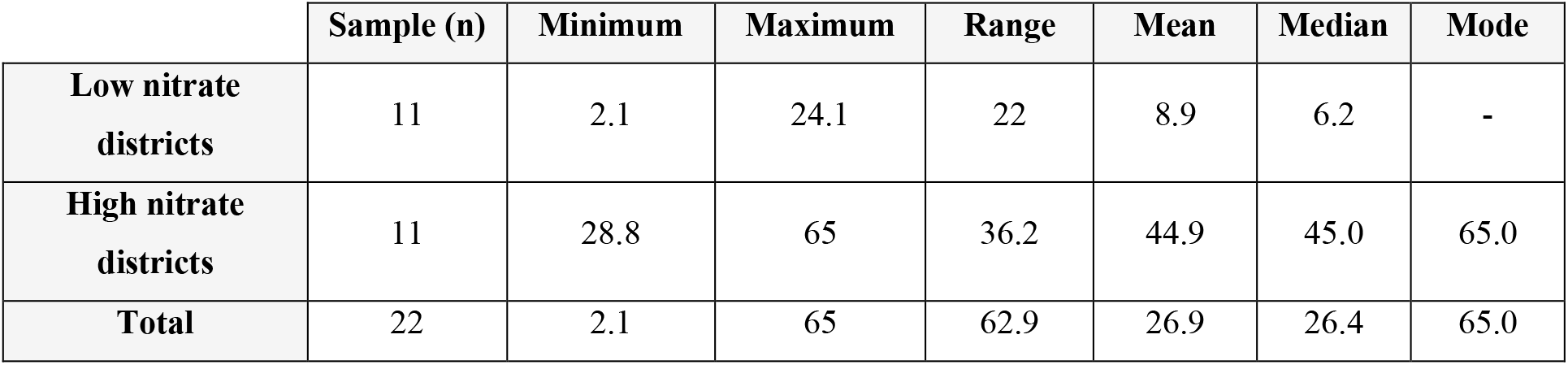
Statistical report of samples.

|  | Sample (n) | Minimum | Maximum | Range | Mean | Median | Mode |
| --- | --- | --- | --- | --- | --- | --- | --- |
| **Low nitrate districts** | 11 | 2.1 | 24.1 | 22 | 8.9 | 6.2 | - |
| **High nitrate districts** | 11 | 28.8 | 65 | 36.2 | 44.9 | 45.0 | 65.0 |
| **Total** | 22 | 2.1 | 65 | 62.9 | 26.9 | 26.4 | 65.0 |

The analysis shows that, based on the Shapiro–Wilk normality test, the distribution of nitrate concentration across the 22 districts of Tehran is not statistically normal (p-value = 0.03) (Figure 5L). Additionally, the comparison results suggest that, at a 95% confidence level, there is some—albeit weak—evidence of a statistically significant difference in mean nitrate concentration among the six Tehran ABFA districts (p-value = 0.06) (Figure 5R).

**Figure 5.**
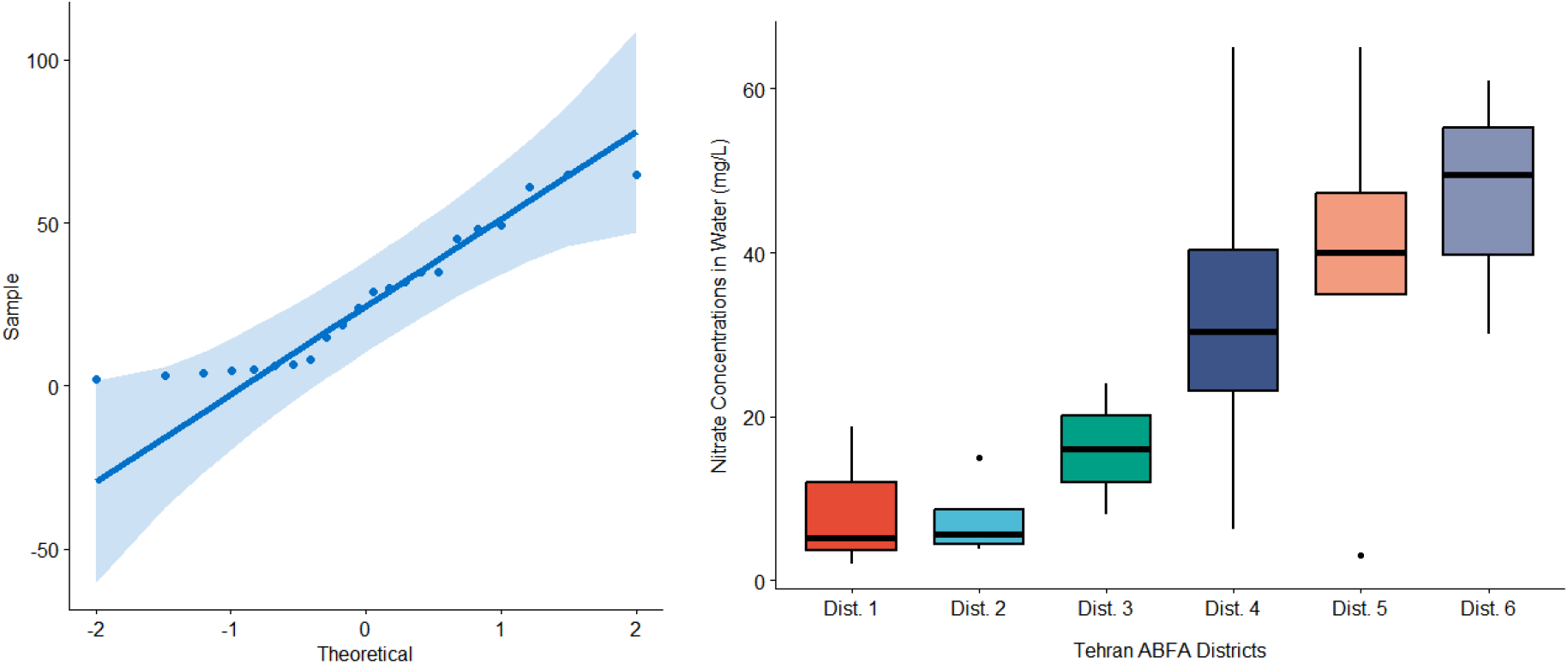
The normality of the distribution of nitrate concentration in the 22 districts of Tehran (left), and Box-plots for the districts of Tehran ABFA districts (right).

The results of HCA on the 22 districts of Tehran are shown in Figure 6.

**Figure 6.**
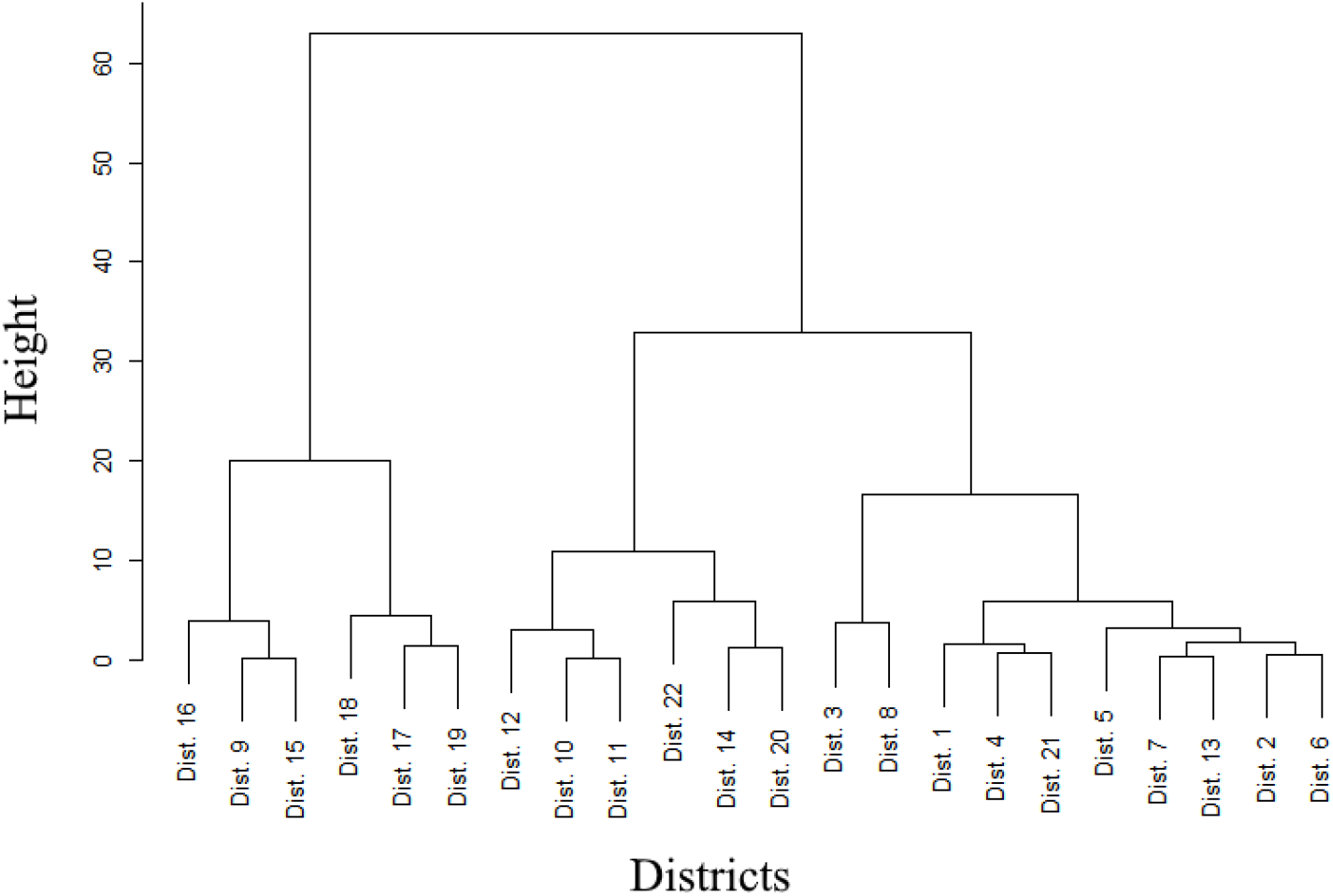
Dendrogram of nitrate distribution in drinking water of 22 districts of Tehran

### 3.2. Relationship Between Social Problems and Nitrate Concentration

Several of the examined social indicators in Tehran were notably higher in districts with elevated nitrate concentrations compared to those with lower levels. These included: the rate of individuals who are divorced or widowed, the percentage of illiterate or low-educated residents, the unemployment rate, perceived nighttime insecurity in neighborhoods, experiences of harassment by vagrants and/or drug addicts, incidents of pickpocketing, physical assaults, feelings of insecurity while alone at home, and the visible presence of addicts and/or vagrants in the neighborhood (Figure 7).

**Figure 7.**
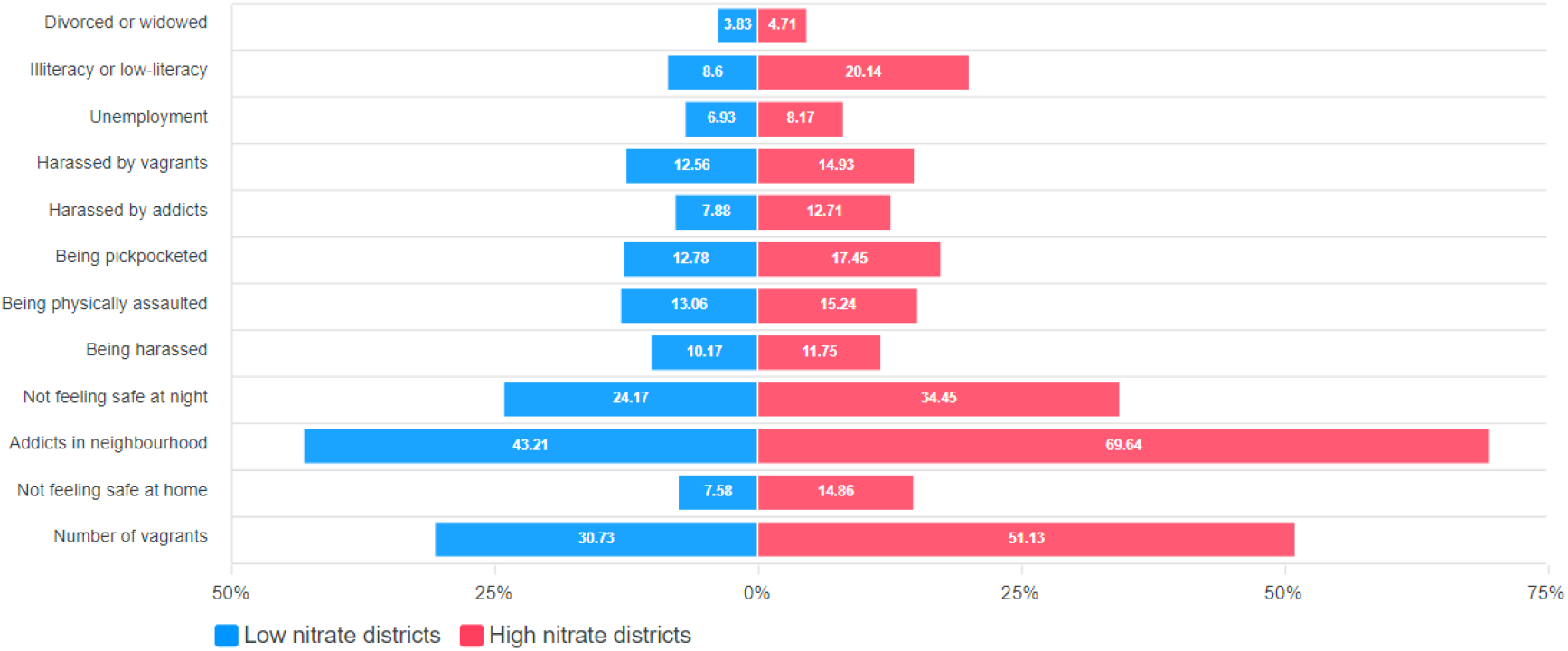
Higher prevalence of some social problems in areas of Tehran that have relatively higher nitrates

In addition, the prevalence percentage of certain social harms in each district showed a direct and significant correlation with the nitrate ion concentration in that district’s drinking water. These included: the percentage of illiterate or low-educated individuals (p-value = 1e-4), harassment by vagrants and addicts (p-value = 0.05 and 4e-4, respectively), the relative ranking of overall social harms (p-value = 0.002), perceived nighttime insecurity in the neighborhood (p-value = 0.02), and the reported number of vagrants and addicts in the neighborhood (p-value = 2e-4 and 3e-4, respectively) (Figure 8).

**Figure 8.**
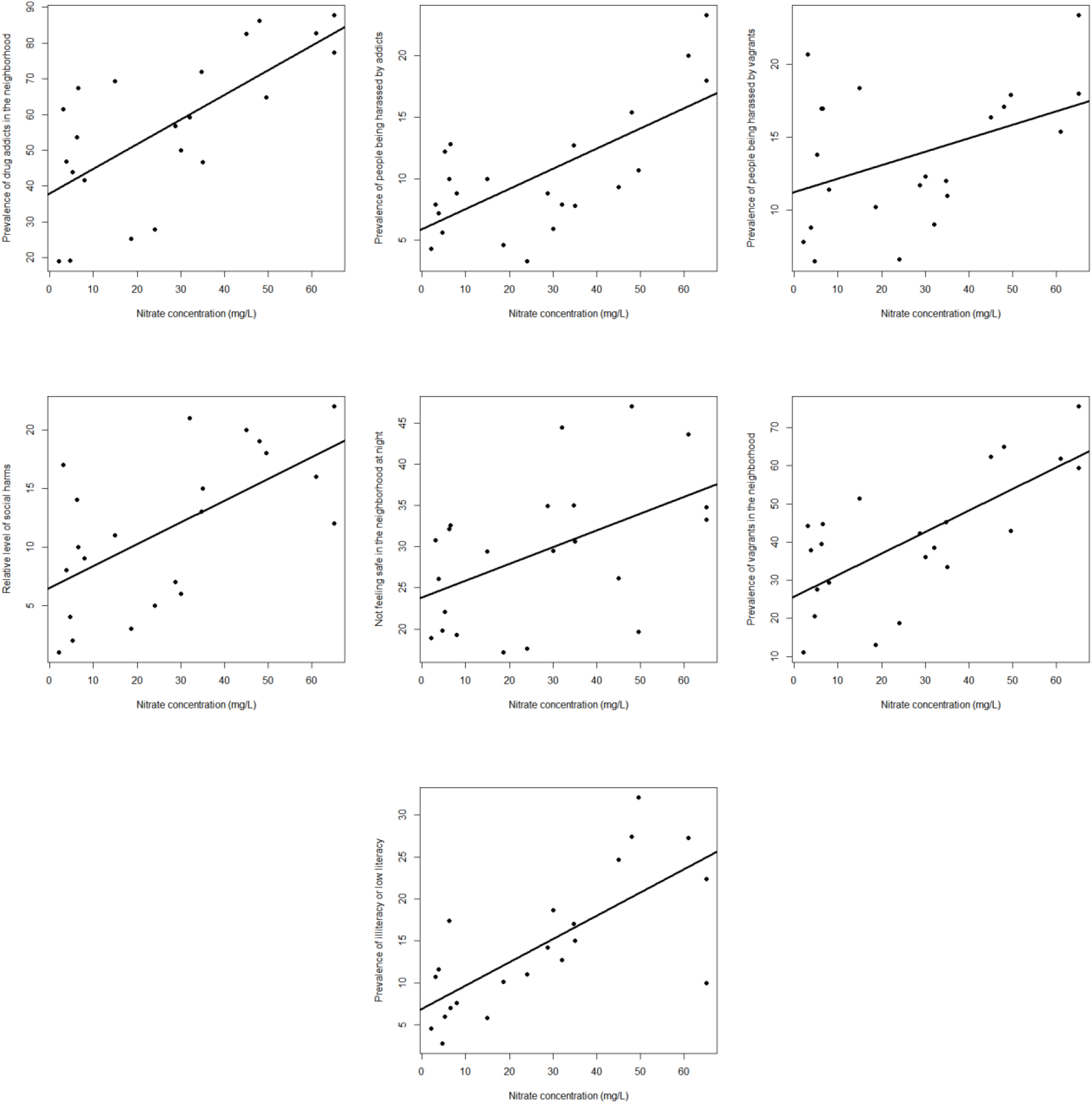
The significant relationship between some social problems and nitrate concentrations in drinking water in 22 districts of Tehran

This is despite the fact that no significant relationship was observed for certain other social harms, such as the rate of burglary and the frequency of bribe requests in municipal offices across districts. Additionally, Figure 9 presents the ranking of districts across several other indicators, with each district assigned a rank from 1 to 22.

**Figure 9.**
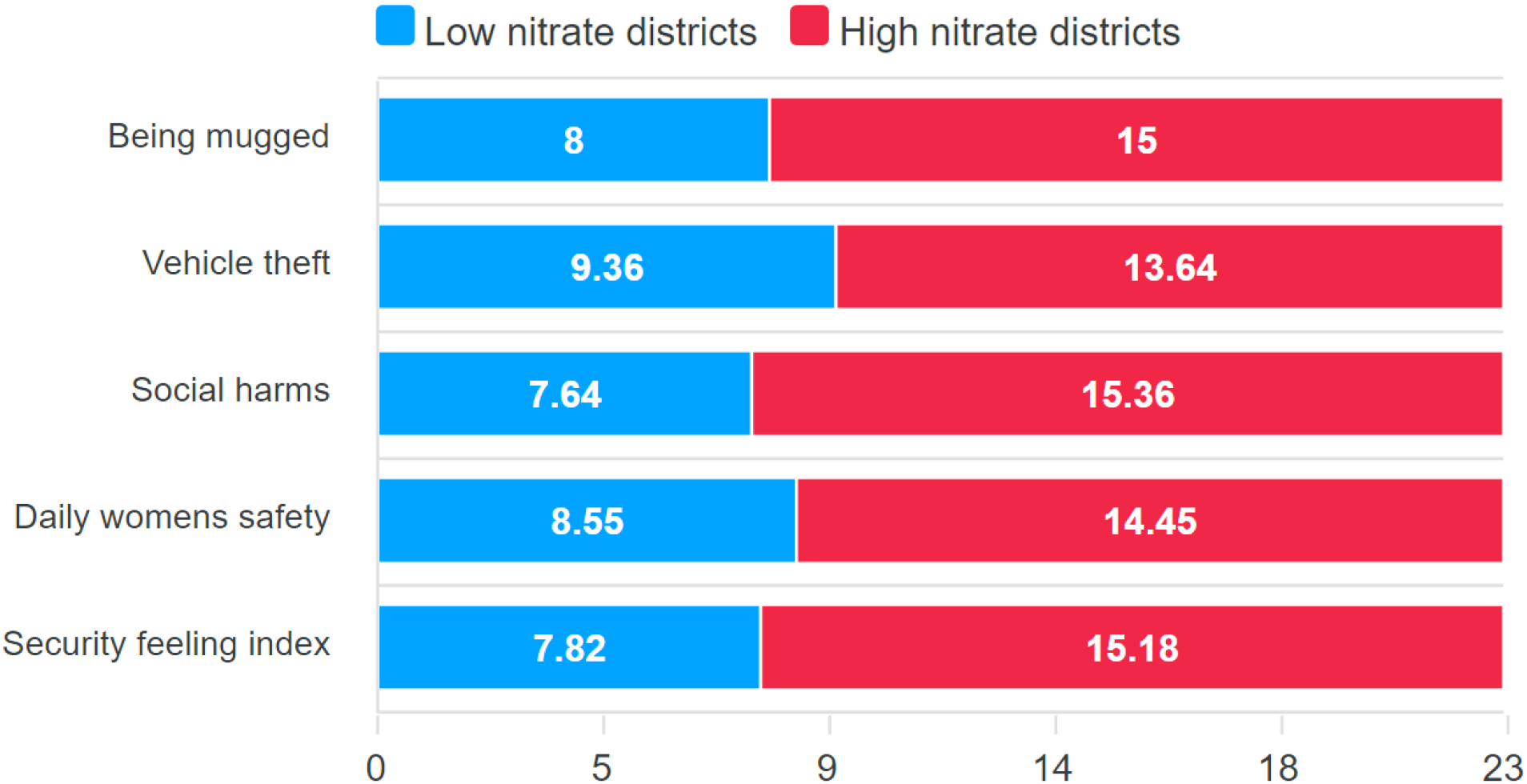
Comparison of the average rank of districts with high nitrates (red) and low nitrates (blue) in various indicators, including ranking of being mugged and vehicle theft, social harms, daily woman safety, and security feeling index.

The findings from this study reveal critical insights into nitrate concentration levels in drinking water across the 22 districts of Tehran, with significant implications for both public health and social conditions. Notably, the elevated nitrate levels in Districts 15, 9, and 16 exceed the Maximum Contaminant Level (MCL) established by the World Health Organization (WHO), raising serious concerns about the safety of drinking water in these areas. The Shapiro–Wilk test indicated a non-normal distribution of nitrate concentrations, suggesting potential influence from outliers or localized factors that require further investigation.

Although the Kruskal–Wallis test showed only weak evidence of significant differences in average nitrate concentrations among Tehran’s six ABFA districts (p-value = 0.06), the result highlights possible spatial variations in water quality. Further research involving a larger sample size or more comprehensive data collection may be necessary to confirm these patterns. These findings emphasize the importance of continuous monitoring and systematic evaluation of drinking water quality across all regions of Tehran to ensure adherence to health standards and minimize the public health risks associated with nitrate exposure.

Moreover, the analysis of social indicators reveals a concerning correlation between elevated nitrate concentrations and various social problems. Districts with higher nitrate levels demonstrated increased rates of divorce, lower educational attainment, and higher unemployment. These associations underscore the potential for environmental factors—such as water quality—to intersect with and intensify existing social challenges. The significant relationships identified between nitrate levels and specific social harms—such as harassment by vagrants and perceived neighborhood insecurity—suggest that communities facing environmental stressors may also be more prone to social instability and vulnerability.

Although some social harms did not show a statistically significant association with nitrate levels, the overall pattern indicates that districts with poorer water quality may be more susceptible to a broader range of social issues. At first glance, this phenomenon may be attributed to multiple factors, including socioeconomic disparities, limited access to education, and unequal distribution of social resources—all of which influence both environmental conditions and social cohesion. This interpretation aligns with findings that impoverished neighborhoods in southern Tehran predominantly rely on well water (Dehkardi, 2022), which is often highly contaminated with nitrates (Asriran, 2018).

Therefore, the evidence points to a correlation—rather than direct causation—between nitrate pollution and the prevalence of social harms in high-nitrate regions. However, a related study conducted by the authors of the present article (Zamaninasab and Bajelan, 2023) examining the relationship between groundwater nitrate levels and social harm prevalence in the United States revealed a comparable trend. In that context, social harms were also clearly correlated with nitrate ion concentrations in groundwater.

## 4. CONCLUSION

The present study highlights the complex and multifaceted relationship between nitrate concentrations in drinking water and a range of social issues affecting Tehran. The findings demonstrate a strong association between elevated nitrate levels and various social harms, a pattern also observed in a companion study examining groundwater quality in the United States. Together, these results provide compelling evidence that the interaction between environmental contaminants and social challenges warrants deeper investigation by researchers across disciplines.

These findings further emphasize the urgent need to integrate environmental and social determinants of health into urban planning and public policy frameworks. As urban areas continue to expand and evolve, understanding how environmental factors—such as drinking water quality—shape social dynamics becomes increasingly critical.

Consequently, future research should focus on conducting longitudinal studies aimed at exploring the potential causal relationship between environmental variables, such as nitrate contamination, and social outcomes. Such studies would enable the assessment of temporal trends and offer clearer insights into causality. Additionally, identifying targeted mitigation strategies is essential for addressing the adverse impacts of high nitrate levels in drinking water, particularly in communities that are disproportionately affected. By focusing on these priorities, future research can support more informed policy decisions and contribute to the development of healthier, more resilient urban environments for all residents.

## Data Availability

All data produced in the present work are contained in the manuscript.

## 5. AUTHOR CONTRIBUTIONS

Z. Bajlan and H. Zamaninasab were responsible for the sampling and analysis of nitrate levels in drinking water across the 22 districts of Tehran. Z. Bajlan and M. Dehghan contributed to the initial drafting of the manuscript. H. Zamaninasab conducted the statistical analyses and prepared the figures. H. Baghari supervised the project, provided critical insights, and reviewed and revised the final version of the article.

## 6. ACKNOWLEDGEMENT

The authors express their sincere gratitude to the staff and officials of Arian Fan Azma Laboratory for their cooperation in the chemical analysis of the water samples.

## 9. ABBREVIATIONS

Not applicable.

